# MRMCsamplesize: An R Package for Estimating Sample Sizes for Multi-Reader Multi-Case Studies

**DOI:** 10.1101/2023.09.25.23296069

**Authors:** Dennis Robert, Saigopal Sathyamurthy, Preetham Putha

## Abstract

Multi-Reader Multi-Case (MRMC) studies are typically used to evaluate improvement in diagnostic accuracy of readers (diagnosticians) when they are assisted by a computer-assisted device (CAD) such as, but not limited to, those based on Artificial Intelligence (AI) algorithms. Statistical analysis of MRMC study data is not trivial and these studies can consume a lot of resources. Optimal planning is crucial and estimation of sample size is a significant step during the study planning phase. MRMC sample size estimations require many parameter assumptions and without pilot data this is generally not intuitive. MRMCsamplesize package can help researchers to estimate sample sizes for an MRMC study in the absence of any pilot data. The program outputs the number of cases required for a given number of readers. The package can also estimate sample sizes for scenarios where intra-cluster correlation (ICC) needs to be adjusted.

## 1. Introduction

Computer-assisted detection (CAD) software devices in healthcare are meant to assist clinicians or diagnosticians (here onwards referred to as ‘readers’) in improving their diagnostic accuracy. Artificial Intelligence/Machine Learning (AI/ML) based CAD devices in the field of Medical AI are becoming increasingly popular. Within the field of Medical AI, the most significant breakthroughs are occurring in the field of Medical Imaging because of a number of reasons, such as the availability of digitized medical images, the image being a representation of the anatomical area of interest in a structured form without much ambiguity as opposed to electronic medical record data and the availability of deep neural networks specifically useful for image pattern recognition such as Convolutional Neural Networks (CNNs). As of August 2023, roughly 500 AI/ML-enabled medical devices have been granted marketing authorization by FDA and approximately 75% of them are intended to be used in the field of Radiology (FDA 2023). A reasonable take on how AI/ML will impact the field of radiology is well captured in the quote, *“Radiologists who use AI will replace those who don’t”*, and not that AI will replace radiologists (Langlotz 2019). Multi-Reader Multi-Case (MRMC) studies are very important in this context because these studies are conducted to test the hypotheses of whether using CAD tools such as those based on AI/ML can improve the diagnostic accuracy of readers. These studies are also accepted by regulatory bodies such as FDA as a means for clinical performance assessment of CAD devices for pre-market notification. (CDRH 2022). MRMC studies are expensive to conduct and consume a lot of resources. The statistical analysis is not trivial due to complex correlation structures. Estimation of a reasonable sample size (both the number of readers and the number of cases) is a crucial element in the study planning phase and this requires making reasonable assumptions of a number of different parameters.

To our knowledge, no R packages are currently available for the estimation of sample sizes for a planned MRMC study without prior pilot data. For example, **RJafroc** (Chakraborty and Zhai 2023) is a comprehensive package to facilitate various types of reader study analysis and it also includes functions to estimate sample sizes. However, it requires pilot data to facilitate input argument assignments, and in the vast majority of scenarios, researchers will have no access to any such pilot data. Even though we can still use functions in **RJafroc** to work without pilot data, this requires the users to supply variance components which are very difficult to conjecture. **MRMCaov** (Smith, Hillis, and Pesce 2023; Smith and Hillis 2020) and **iMRMC** (Gallas 2023) are both useful for the statistical analysis of MRMC study data, but they do not contain functions for the estimation of sample sizes to aid study planning. There are JAVA-based software programs available for sizing an MRMC study. One example is an open-source JAVA-based graphical user interface (GUI) program (Hillis and Schartz 2018) developed at the University of Iowa. This program allows users to perform sample size estimation for MRMC studies and it can do so for a wide variety of options such as with and without pilot data, various types of study designs such as random readers and random cases, fixed readers and fixed cases, etc., different methods such as both OR and DBM methods. Another JAVA-based GUI program named *iMRMC* (Gallas, Pennello, and Myers 2007), developed by the same research team who developed the **iMRMC** package, is also available for sample size estimations. Without pilot data, it might not be intuitive to supply variance components or correlation components which are required as inputs for sample size estimations in these programs. In addition, intra-cluster correlation (ICC) due to the presence of multiple target lesions within a diseased case (for example, the presence of multiple lung nodules in a chest X-ray image) further complicates the analysis because the diagnostic accuracy is dependent not just on detecting a lesion *somewhere* in a case, but also localizing it correctly. In such scenarios, sample size estimations have to be adjusted for the anticipated ICC in diseased cases. Throughout this article we use the term “diseased cases” for cases with target lesion of interest and “non-diseased cases” for cases without any target lesion of interest.

**[MRMCsamplesize](https://cran.r-project.org/package=MRMCsamplesize)** (Robert 2023), the R package of focus in this article, can estimate the number of required cases for a given number of readers for a fully-crossed MRMC study using the Obuchowski-Rockette (OR) method (Nancy A. Obuchowski and Rockette 1995). It can be used for sizing MRMC studies with or without adjustment for anticipated ICC. The sample size output from **[MRMCsample-size](https://cran.r-project.org/package=MRMCsamplesize)** was validated by comparing it against published literature. The results of this validation exercise are also presented in this article.

## 2. Fundamentals of MRMC Study Design and Statistical Analysis

In an MRMC study, a set of readers interpret a set of cases (typically medical images) with and without AI. A hypothetical example of a fully-crossed MRMC study workflow with a single reader and five images is illustrated in Figure 1.

**Figure 1:**
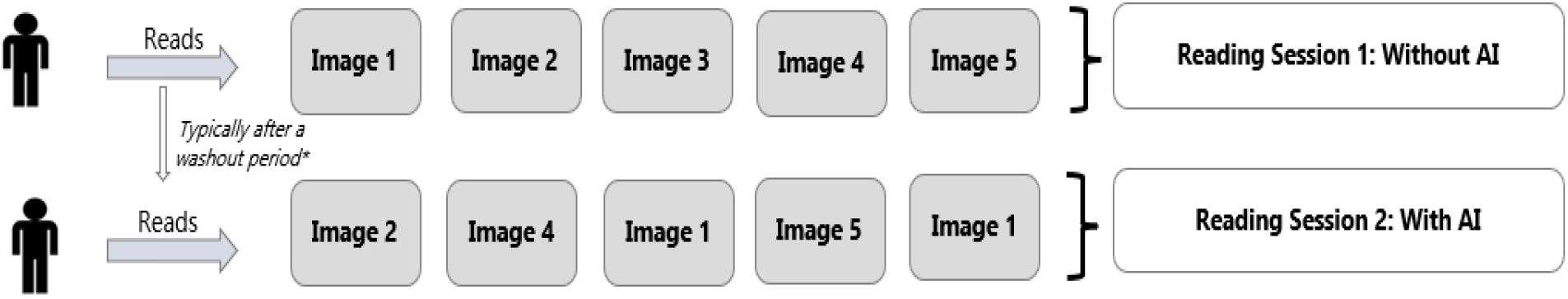
A hypothetical example of an MRMC study workflow with a single reader and five images. The order in which the reader reads the images is typically randomized in each session to mitigate reading-order bias. *Washout period of 4 weeks is typically used in cross-over designs, but sequential designs without any washout period may also be used depending on the intended use of the CAD/AI device. It is also not rare to mix reading sessions with cases from both modalities. For example, Reading Session 1 can include a mix of images with and without the assistance of AI and similarly for Reading Session 2 as well. In such scenarios, if an image is read by a reader with AI assistance in the first session, then in the second session, that image has to be read by that reader without AI assistance.

Since the same reader reads the same image twice and the same images are read by different readers, there are intra- and inter-reader variabilities which result in complex correlation that needs to be considered during statistical analysis. The choice of whether the design has a sequential nature or cross-over nature can also have an impact on sample size estimations (Nancy A. Obuchowski and Bullen 2022).

While multiple different statistical methods are proposed for the analysis of MRMC study data, two methods are particularly popular. They are:

- Obuchowsiki-Rockette (OR) method
- Dorfman-Berbaum-Metz (DBM) method

The OR method fits a correlated-by-error-test-by-reader ANOVA (a mixed effects ANOVA model), treating readers as random effects, the interaction between test and reader as another random effect and a fixed effect for the test as independent variables and average per-test reader performance outcomes such as area under the receiver operating characteristics curve (AUC) as the dependent variable. (Nancy A. Obuchowski and Rockette 1995). The DBM method fits a test-by-reader-by-case conventional ANOVA to case-specific pseudovalues (Dorfman, Berbaum, and Metz 1992). OR method is considered to be more intuitive since the parameters are more interpretable due to the method modelling the observed reader-performance outcomes rather than the pseudovalues (Iowa 2023). **[MRMCsamplesize](https://cran.r-project.org/package=MRMCsamplesize)** package is developed based on the OR method.

## 3. Sample Size Estimation for MRMC Studies

### 3.1. Endpoints

The primary endpoints for MRMC studies are typically the difference in reader performance as quantified by a figure-of-merit (FOM) such as AUC, sensitivity (Se), specificity (Sp) or area under the free-response curve (FROC) paradigm such alternative-FROC (AFROC).

**[MRMCsamplesize](https://cran.r-project.org/package=MRMCsamplesize)** can estimate sample size if the planned primary endpoint is difference in AUC or Se. The methodology of sample size estimation is detailed in the next section.

### 3.2. Detailed Methodology

It has to be known during the study planning whether the diseased cases can contain multiple target lesions of interest or not. For example, multiple lung nodules can be present in a single chest X-ray image and the reader’s accuracies are evaluated by the number of correct lesions that the reader identifies. In this case, there can be correlations between the lesions in the same diseased case, and the resulting *ICC* has to be thus adjusted. The first step is to estimate a required number of diseased cases (cases with target lesions of interest) and non-diseased cases (cases with no target lesions of interest) assuming independence (i.e., no adjustment for *ICC*) and then adjust the numbers by accounting for the design effect (*DE*) resulting from the anticipated *ICC* (Nancy A. Obuchowski and Hillis 2011).

#### 3.2.1. Assuming independence between observations

The methods described here are largely based on the methods published by Obuchowski, Rockette and Hillis in multiple publications (Nancy A. Obuchowski and Rockette 1995; Nancy A. Obuchowski 2000; Hillis and Schartz 2018; Zhou, Obuchowski, and McClish 2011). The words ‘case’ and ‘image’ are used interchangeably in this article.

Assuming that the design is fully-crossed and that there are two modalities to be studied (for example, each reader reads each case twice, one with AI and one without AI), the null and alternative hypothesis is:

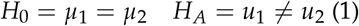

In (1), *μ*_*i*_ is the mean FOM of readers for the modality i.

For the sample size estimation, under the alternative hypothesis, it is specified that *μ*_1_ − *μ*_2_ = Δ, where Δ is the suspected difference (effect size) in the FOM of interest. Obuchowski and Rockette proposed a modified F statistic to test the null hypothesis using a 2-way mixed effects ANOVA model (Nancy A. Obuchowski and Rockette 1995). The noncentrality parameter of the F-distribution, denoted by *λ*, can be used to derive sample size estimates with pre-specified power and type 1 error rate to be able to detect a pre-specified minimum effect size of Δ. *λ* is given by:

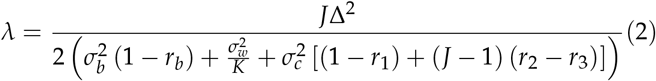

The definitions of the parameters in the RHS (Right-Hand Side) of equation (2) are listed in Table 1.

**Table 1:**
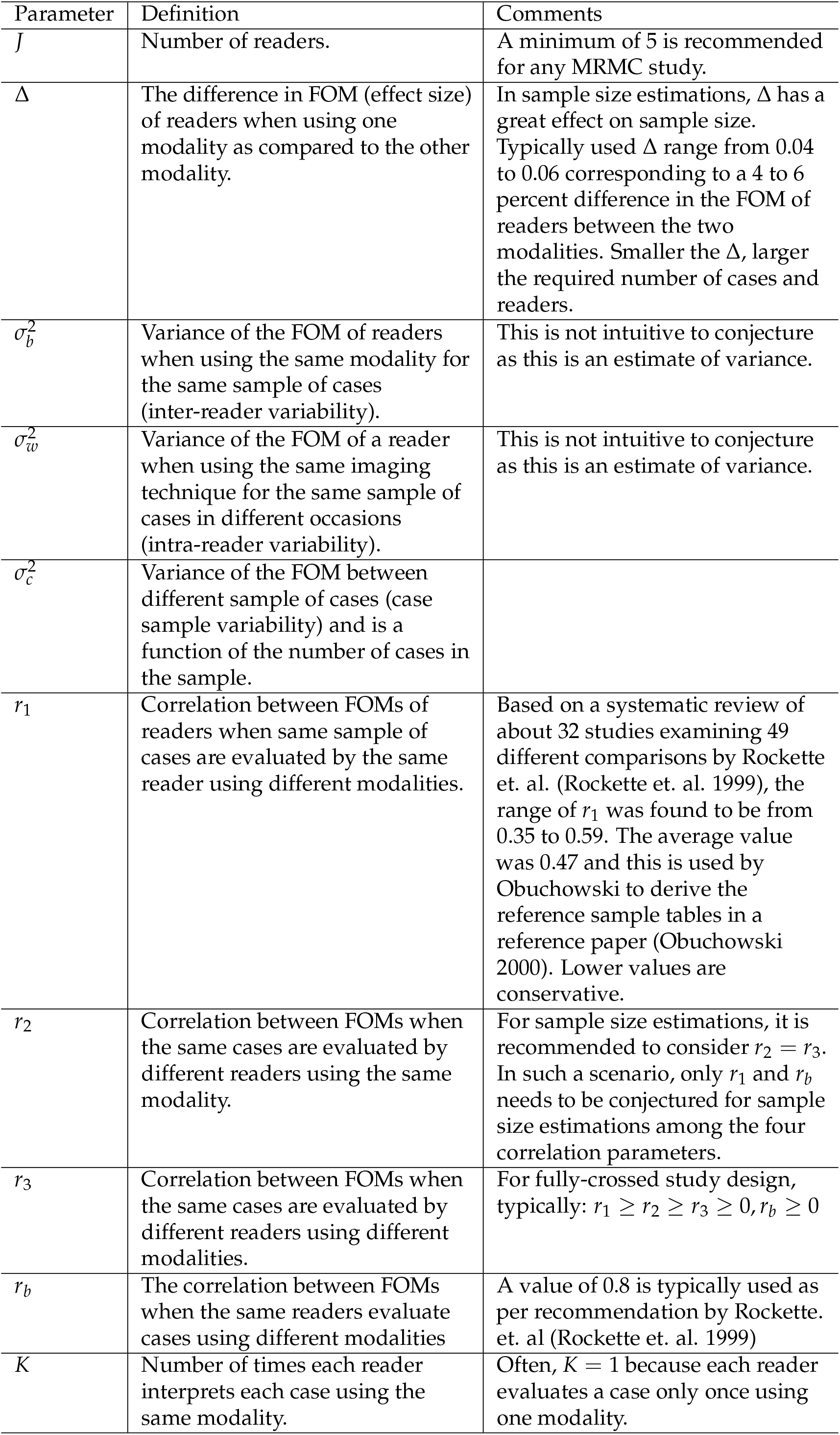
Parameters and definitions.

The estimated power *p* of a study with J readers is:

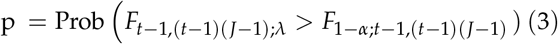

Where *F*_*t*−1,(*t*−1)(*J*−1);*λ*_ denotes a random variable having a non-central F distribution with degrees of freedom *t* − 1 and (*t* − 1)(*J* − 1) and non-centrality parameter of *λ, t* is the number of modalities used in the study and *F*_1 − *α*;*t* − 1,(*t* − 1)(*J* − 1)_ is the (1 − *α*) 100*th* percentile of a central F distribution with the same degrees of freedom.

The non-centrality parameter of F distribution, *λ*, that would provide a pre-specified power *p* and type I error rate *α* can be calculated. This calculation needs numerator and denominator degrees of freedom also to be pre-specified in addition to required *p* (typically 0.8) and *α* (typically 0.05). The numerator and denominator degrees of freedom used for calculating *λ* are respectively *t* − 1 and (*t* − 1)(*J* − 1) (Zhou, Obuchowski, and McClish 2011). For example, *λ* that would provide 80% power with a type 1 error rate of 5% are 18.12, 12.36, 9.92 and 8.72 when J is equal to 4, 6, 10 and 20, respectively.

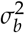 and 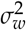 are difficult to conjecture in the absence of pilot data but are very important parameters that determine the power of the study. Conjecturing the range of FOMs is much more intuitive. If the FOMs of the readers are assumed to follow a normal distribution, the relationship between range and standard deviation can be used to derive the variances 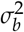 and 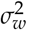 by multiplying the conjectured range by a constant derived from normal distribution (Nancy A. Obuchowski 2000). Another alternative approach is to use the simple rule that standard deviation equals the range divided by four as approximately 95% of the values are distributed within four standard deviations. The former method is more conservative, especially when the number of readers is less than 30-35, and thus may be the more safer approach. This is the default method used in **[MRMCsamplesize](https://cran.r-project.org/package=MRMCsamplesize)** to derive inter- and intra-reader variances from the intra- and inter-reader variability ranges conjectured.

The only remaining unknown parameter in equation (2) is 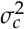 and this can be computed by rearranging equation 3. 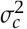 is in turn, a function of the anticipated average FOM (AUC or Se). In the case when the FOM of interest is AUC, 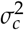 is given by:

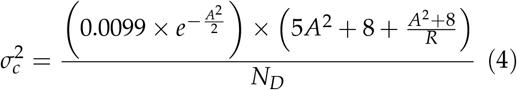

where *A* is:

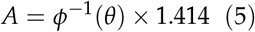

In equation (5), *θ* is the anticipated average AUC of readers and *ϕ*^−1^(*θ*) is the inverse cumulative normal distribution function. *R* is the ratio of non-diseased cases to diseased cases and *N*_*D*_is the number of diseased cases. Note that equations (4) and (5) are based on estimating variance of AUC assuming a binormal distribution as described by Zhou, Obuchowski and McClish (Zhou, Obuchowski, and McClish 2011). Blume (Blume 2009) recommended another approach to estimate the variance of AUC which requires no parametric assumptions. This is also implemented in the **[MRMCsamplesize](https://cran.r-project.org/package=MRMCsamplesize)** package.

When the FOM is sensitivity (Se), 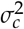 can be estimated by considering the properties of a binomial proportion estimate and its variance as given by:

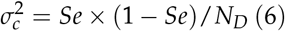

From here onwards, the equations will be based on the assumption that the FOM of interest is AUC. For Se, the only difference will be re-arrange the equations (2) and (6). Using (2) and (4), the number of diseased cases required for the MRMC study can be estimated by:

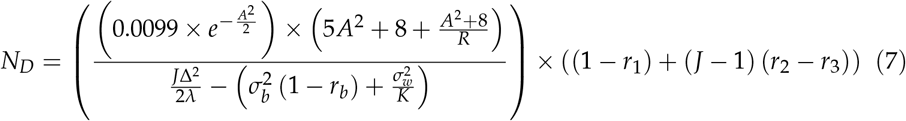

Note that in (7), the term 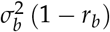 is also known as the *test* − *by* − *reader* − *variance* in some literature and it is the variance of the interaction between the modality and the reader (Nancy A. Obuchowski and Hillis 2011).

The total sample size (sum of diseased and non-diseased cases), *N*_*T*_required for the planned study would thus be:

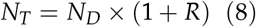

##### 3.2.1.1. Estimating inter- and intra-reader variances using range

One can employ two methods for deriving 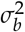 and 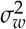 using corresponding ranges, *range*_*b*_ and *range*_*w*_, respectively, the latter two being much more intuitive to conjecture in the absence of any pilot data.

*range*_*b*_: The anticipated difference between the highest accurate (highest FOM) reader in the study and the lowest accurate (lowest FOM) reader.

*range*_*w*_: The anticipated difference between the FOMs of a reader who interprets the same cases using the same modality at two different times.

The first method assumes that readers’ FOMs follow a normal distribution and thus *σ*_*b*_ and *σ*_*w*_can be computed by multiplying *range*_*b*_ and *range*_*w*_by constants *c*_*b*_ and *c*_*w*_derived from the normal distribution. Note that *c*_*b*_ depends on the number of readers, but *c*_*w*_is dependent on the number of modalities used in the study (typically, there are two modalities or reading sessions per reader). Three statistical properties are used for the estimation of *c*_*b*_ and *c*_*w*_:

- The expected range for a sample of size *n* in a symmetric distribution with mean 0 is twice the expected largest value in a sample of the same size.
- The density of the largest value *X*_*n*_in a sample of size *n* from a distribution with density *f* and cumulative distribution function *F* is *n × f* (*x*) *× F*(*x*)^*n*−1^
- For a normal distribution sample with size *n*, the expected range is the expected range of a standard normal sample of size *n* times the standard deviation.

So the expected largest value *X*_*n*_ in a sample of size *n* following a nozrmal distribution is obtainable by integration as per order statistics.

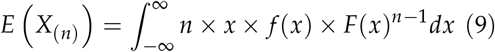

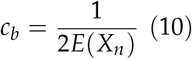

As an example, when J is 10 and *range*_*b*_ is assumed to be 0.1 (10% difference in FOM between the highest and lowest accurate readers), *σ*_*b*_ is calculated by:

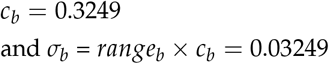

*c*_*w*_, as discussed above, is dependent only on the number of reader sessions per reader and this is typically always 2 in a fully crossed MRMC study assessing accuracies of two modalities. Hence *c*_*w*_is 0.8862 in most scenarios regardless of J. The calculation of *sigma*_*w*_from *c*_*w*_is trivial.

The second method is rather trivial and uses the relationship between range and standard deviation.

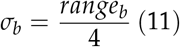

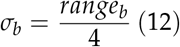

Note that in order to get the variances 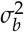 and 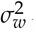, the standard deviations *σ*_*b*_ and *σ*_*w*_must be squared.

Between the two methods, first method is the more conservative method especially when number of readers (*J*) is less than 30.

### 3.2.2. Adjusting for intra-cluster correlation

For taking into account the fact that there can be multiple target lesions in a single case and it is required to evaluate the accuracy of readers based on detecting all the lesions in all the diseased cases, one must conjecture an anticipated *ICC* and the average number of target lesions in diseased cases (*s*). The design effect (*DE*) is a function of *ICC* and *s*. This will yield the 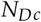 which is the number of diseased cases after adjustment for the *ICC*. Note that *N*_*Dc*_is always less than or equal to 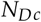 and this means that number of diseased cases required for a MRMC study where *ICC* is expected is less than the number that is required where this adjustment is not required given the same assumptions for all other parameters. An *ICC* of 0.5 is often used and is a moderately conservative. *s* is often derived from literature review or is assumed conservatively.

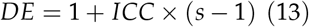

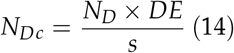

## 4. Using MRMCsamplesize

The function within the **[MRMCsamplesize](https://cran.r-project.org/web/packages/MRMCsamplesize/index.html)** package that can be used for estimating sample sizes for fully-crossed MRMC studies is *sampleSize*_*MRMC*. Details of the arguments of this function are detailed in Table 2.

**Table 2:** Function arguments and definitions.

| Argument | Definition |
| --- | --- |
| *endpoint* | Character string to inform what is the endpoint of the MRMC study. Values can be either *auc* or *sensitivity* |
| *J* | The number of readers for the study. It is recommended to have minimum 5 readers in any MRMC study. |
| *delta* | Effect size denoting the anticipated difference in the endpoint between the two interventions/imaging-modalities/techniques. Typically chosen values are 0.04, 0.05 and 0.06. Should be between 0 and 1. |
| *rangeb* | Inter-reader variability (sometimes referred to as between-reader variability) range denoting the anticipated difference between the highest accuracy of any reader in the study and the lowest accuracy of any reader in the study. Should be a numeric value between 0 and 1.). |
| *rangew* | Intra-reader variability range (sometimes referred to as within-reader variability) denoting the anticipated difference between the accuracies of a reader who interprets the same images using the same imaging technique at two different times. Should be a numeric value between 0 and 1. |
| *theta* | Expected average value of the endpoint for the J readers. Should be a numeric value between 0 and 1. |
| *R* | Ratio of non-diseased cases to diseased cases. Defaults to 1. |
| *r1* | Correlation between FOMs of readers when same cases are evaluated by the same reader using different modalities. |
| *r2* | Correlation between FOMs when the same cases are evaluated by different readers using the same modality. It is assumed that $r2 = r3$ for default calculations |
| *r3* | Correlation between FOMs when the same cases are evaluated by different readers using different modalities. It is assumed that $r2 = r3$ for default calculations. |
| *rb* | Correlation between FOMs when the same readers evaluate cases using different modalities. Defaults to 0.8. |
| *K* | Number of times each reader interprets the same images from the same modality. $K = 1$ in a fully-crossed paired-reader paired-case study design with two modalities. |
| *power* | Power to detect delta given all other assumptions. Default value is 0.8 corresponding to 80 percent power. |
| *alpha* | The type I error rate. Default value is 0.05 corresponding to 5 percent type I error (significance level). |
| *nu1* | Numerator degrees of freedom of the F-distribution which will be used to estimate the non-centrality parameter. Defaults to 1. |
| *var_auc* | Variance estimation method when endpoint is *auc*. Defaults to *obuchowski* based on Zhou et.al. (2011). If value is changed to *blume*, then method proposed by Blume (2009) will be used to estimate the variance. |
| *reader_var_estimation_method* | Method to be used to estimate inter- and intra-reader variances from *rangeb* and *rangew*. Defaults to *normal* which corresponds to the first method in section 3.2.2.1. |
| *n_reading_sessions_per_reader* | Number of times each reader interprets each case. Defaults to 2 which corresponds to a typical MRMC study with 2 modalities. |
| *corr* | Logical value indicating if ICC has to be adjusted or not. Defaults to *FALSE* which indicates adjustment is not required. If *TRUE*, then both *ICC* and *s* have to be specified as valid numerical values. |
| *ICC* | A numerical value between 0 and 1 indicating the expected ICC. |
| *s* | Average number of lesions in diseased cases. Should be a numeric value which is more than 1. |

### 4.1. Example 1: Without adjustment for ICC

#### Scenario

An AI software to detect and localize adenomas from CT colonography scans is available. Will using this device increase the accuracy (in terms of AUC) of radiologists (readers) in detecting adenomas in CT colonography? Only case-level AUCs are to be estimated and correlation arising from the presence of adenomas in a single scan can be ignored. Determine a sample size for an MRMC study considering readers with varying experience (general radiologists, radiologists with abdominal radiology fellowships, 1-12 yrs experience)

#### Sample size estimation result

sample of 20 readers and 436 scans (218 with presence of adenomas, 218 with no adenomas) will have 80% power at a type I error rate of 5% to detect a minimum difference in readers’ AUC of 5% assuming a large inter-reader and intra-reader variability of 20% and 5% respectively, a 0.47 moderate correlation between readers, anticipated average readers’ AUC as 0.75.

The programmatic implementation of sample size estimation for the example is shown below:

~~~
ex1 <- sampleSize_MRMC(endpoint = ’auc’,
 J = 20,
 delta = 0.05,
 theta = 0.75,
 r1 =0.47,
 rangeb = 0.20,
 rangew = 0.05)
ex1$ORSampleSizeResults
#>
#> Obuchowski-Rockette Sample Size Estimation Results
#>
#> ICC = Not applicable
#> nUnits_i = 218
#> nCases_c = NA
#> nControls = 218
#> nTotal = 436
#> J = 20
#> DE = NA
#> s = NA
#> power = 0.8
#> alpha = 0.05 #>
#> NOTE:
#> ICC: Is intra-cluster correlation (ICC) considered while estimating sample size?
#> nUnits_i: Number of required units (a unit is a lesion of interest) assuming independence between units
#> nCases_c: Number of required diseased cases with presence of at least one unit of lesion after adjusting for ICC
#> nControls: Number of required non-diseased cases
#> nTotal: Total sample size (cases) #> J: Number of readers
#> DE: Design effect due to ICC
#> s: Assumed average number of lesions in diseased cases
#> power: Assumed power
#> alpha: Significance level (Type I error rate)
~~~

### 4.2. Example 2: With adjustment for ICC

#### Scenario

A single CT colonography scan can have multiple adenomas. The scenario is same as in example 1, but with adjustment for correlation is needed. Not only that the radiologist need to detect a lesion (s), they also have to localize the detected lesion (s) correctly. The task is to determine a sample size for an MRMC study considering readers with varying experience (general radiologists, radiologists with abdominal radiology fellowships, 1-12 yrs experience) with primary endpoint being difference in AUC of readers when aided by the AI device as compared to when not aided by it.

#### Sample size estimation result

A sample of 20 readers and 394 scans (197 with presence of at least one adenoma, 197 with no adenoma) will have 80% power at a type I error rate of 5% to detect a minimum difference in readers’ AUC of 5% assuming a large inter-reader and intra-reader variability of 20% and 5% respectively, a 0.47 moderate correlation between readers, anticipated average readers’ AUC as 0.75, a moderate ICC of 0.5 due to the presence of multiple nodules in a single scan and average number of adenomas in scans with adenomas as 1.25.

~~~
ex2 <- sampleSize_MRMC(endpoint = ’auc’,
 J = 20,
 delta = 0.05,
 theta = 0.75,
 r1 =0.47,
 rangeb = 0.20,
 rangew = 0.05,
 corr = TRUE,
 ICC = 0.5,
 s = 1.25)
print(ex2$ORSampleSizeResults)
#>
#> Obuchowski-Rockette Sample Size Estimation Results
#>
#> ICC = Intra-class correlation applicable
#> nUnits_i = 218
#> nCases_c = 197
#> nControls = 197
#> nTotal = 394
#> J = 20
#> DE = 1.125
#> s = 1.25
#> power = 0.8
#> alpha = 0.05
#>
#> NOTE:
#> ICC: Is intra-cluster correlation (ICC) considered while estimating sample size?
#> nUnits_i: Number of required units (a unit is a lesion of interest) assuming independence between units
#> nCases_c: Number of required diseased cases with presence of at least one unit of lesion after adjusting for ICC
#> nControls: Number of required non-diseased cases
#> nTotal: Total sample size (cases)
#> J: Number of readers
#> DE: Design effect due to ICC
#> s: Assumed average number of lesions in diseased cases
#> power: Assumed power
#> alpha: Significance level (Type I error rate)
~~~

## 5. Validation

In order to validate the results from **[MRMCsamplesize](https://cran.r-project.org/package=MRMCsamplesize)**, the sample size numbers from published literature (Nancy A. Obuchowski 2000) were used. The results from **[MRMCsamplesize](https://cran.r-project.org/web/packages/MRMCsamplesize/index.html)** was compared against this published literature. A total of 162 MRMC design scenarios were simulated by varying the number of readers, inter- and intra-reader variability ranges, effect size (delta), ratio of non-diseased to diseased cases and anticipated average AUC by assuming independence (without any adjustment for ICC). The results from the R function were found to be comparable to the numbers from a published reference. Of the 162 simulations, 11 sample size outputs from R (7%) were moderately different from that of the reference value (overestimated by >5% or underestimated by more than 1%). The maximum absolute percentage deviation was 5.26% from the reference number. Most of the differences were found in the small number of reader scenarios (J=4) when assumed reader variabilities are large. Such a scenario is usually not advised anyway as a minimum of five readers is typically needed to get a reliable estimate of the variances in the accuracy of readers. The tables listing the results from this validation exercise are available in the supplementary material (S1).

## Data Availability

There is no specific data used for this research. The validation results can be reproduced using the R script available online at: https://github.com/technOslerphile/MRMCsamplesize/blob/master/S1.R

## 6. Acknowledgements

The authors want to acknowledge and thank the colleagues at Qure.ai for their support and review of the manuscript. Many existing reference literature provided the much-needed inputs to this work which would have been impossible in the absence of such literature. In that regard, we want to especially acknowledge the authors of the original Obuchowski-Rockette model.

