## Supplementary material for "MRMCsamplesize: An R Package for Estimating Sample Sizes for Multi-Reader Multi-Case Studies": S1

### Validation Results

- Small, moderate and large reader variabilities correspond to variations in intra- and inter-reader variability ranges and were defined as per the definitions in the reference literature (Obuchowski 2000).

| Variability | Inter-reader variability range, $range_b$ | Intra-reader variability range, $range_w$ |
|---|---|---|
| Small | 0.01 | 0.005 |
| Moderate | 0.05 | 0.025 |
| Large | 0.10 | 0.05 |

- Table 1, Table 2 and Table 3 contains the results when number of readers is assumed to be 4, 6 and 10 respectively in the MRMC study. Within each table, a different number of scenarios were modelled by varying AUC, effect size and ratio of non-diseased to diseased cases.
- The *'Reference Sample Size'* field in the table shows the number of cases from the reference literature and the *'R Output Sample Size'* shows the number of cases obtained from the *MRMCsamplesize* package together with percentage difference from the reference sample size.
- All calculations assumed a power of 80% and a type 1 error rate of 5%.
- When the number of diseased cases required turned out to be <10, a minimum of 10 diseased cases was assigned to that scenario because it is not recommended to conduct an MRMC study with anything less than that.
- An overestimation by >5% or underestimation by >1% are coloured red.
- The R script to reproduce the 'R output Sample Size' estimates in Table 1, Table 2 and Table 3 are available here: https://github.com/technOslerphile/MRMCsamplesize/blob/master/S1.R

**Table 1**

| | | | Number of Readers (J) = 4 | | | | | |
|---|---|---|---|---|---|---|---|---|
| | | | Small Reader Variability | | Moderate Reader Variability | | Large Reader Variability | |
| AUC (theta) | Effect Size (delta) | Ratio (R) | Reference Sample Size | R Output Sample Size (% Diff) | Reference Sample Size | R Output Sample Size (% Diff) | Reference Sample Size | R Output Sample Size (% Diff) |
| 0.75 | 5% | 1 | 571 | 568 (-0.53%) | | | | |
| | | 2 | 679 | 675 (-0.59%) | | | | |
| | | 4 | 983 | 980 (-0.31%) | | | | |
| | 10% | 1 | 133 | 134 (0.75%) | 291 | 290 (-0.34%) | | |
| | | 2 | 159 | 159 (0%) | 345 | 345 (0%) | | |
| | | 4 | 229 | 230 (0.44%) | 500 | 500 (0%) | | |
| | 15% | 1 | 59 | 60 (1.69%) | 77 | 78 (1.3%) | 2975 | 2912 (-2.12%) |
| | | 2 | 70 | 72 (2.86%) | 92 | 93 (1.09%) | 3536 | 3462 (-2.09%) |
| | | 4 | 101 | 100 (-0.99%) | 132 | 135 (2.27%) | 5122 | 5015 (-2.09%) |
| 0.90 | 5% | 1 | 287 | 290 (1.05%) | | | | |
| | | 2 | 363 | 366 (0.83%) | | | | |
| | | 4 | 548 | 550 (0.36%) | | | | |
| | 10% | 1 | 67 | 68 (1.49%) | 146 | 148 (1.37%) | | |
| | | 2 | 85 | 87 (2.35%) | 185 | 186 (0.54%) | | |
| | | 4 | 128 | 130 (1.56%) | 279 | 280 (0.36%) | | |
| | 15% | 1 | 30 | 30 (0%) | 39 | 40 (2.56%) | 1497 | 1478 (-1.27%) |
| | | 2 | 38 | 39 (2.63%) | 49 | 51 (4.08%) | 1890 | 1869 (-1.11%) |
| | | 4 | 57 | 60 (5.26%) | 74 | 75 (1.35%) | 2856 | 2820 (-1.26%) |

**Table 2**

| | | | Number of Readers (J) = 6 | | | | | |
|---|---|---|---|---|---|---|---|---|
| | | | Small Reader Variability | | Moderate Reader Variability | | Large Reader Variability | |
| AUC (theta) | Effect Size (delta) | Ratio (R) | Reference Sample Size | R Output Sample Size (% Diff) | Reference Sample Size | R Output Sample Size (% Diff) | Reference Sample Size | R Output Sample Size (% Diff) |
| 0.75 | 5% | 1 | 246 | 246 (0%) | 3769 | 3694 (-1.99%) | | |
| | | 2 | 293 | 291 (-0.68%) | 4479 | 4392 (-1.94%) | | |
| | | 4 | 424 | 425 (0.24%) | 6488 | 6360 (-1.97%) | | |
| | 10% | 1 | 60 | 60 (0%) | 78 | 78 (0%) | 943 | 924 (-2.01%) |
| | | 2 | 71 | 72 (1.41%) | 92 | 93 (1.09%) | 1120 | 1098 (-1.96%) |
| | | 4 | 103 | 105 (1.94%) | 133 | 135 (1.5%) | 1622 | 1590 (-1.97%) |
| | 15% | 1 | 27 | 28 (3.7%) | 30 | 30 (0%) | 46 | 46 (0%) |
| | | 2 | 32 | 33 (3.13%) | 35 | 36 (2.86%) | 54 | 54 (0%) |
| | | 4 | 50 | 50 (0%) | 51 | 55 (7.84%) | 78 | 80 (2.56%) |
| 0.90 | 5% | 1 | 124 | 126 (1.61%) | 1896 | 1876 (-1.05%) | | |
| | | 2 | 157 | 159 (1.27%) | 2395 | 2370 (-1.04%) | | |
| | | 4 | 236 | 240 (1.69%) | 3618 | 3580 (-1.05%) | | |
| | 10% | 1 | 31 | 32 (3.23%) | 39 | 40 (2.56%) | 474 | 470 (-0.84%) |
| | | 2 | 38 | 39 (2.63%) | 50 | 51 (2%) | 599 | 594 (-0.83%) |
| | | 4 | 58 | 60 (3.45%) | 75 | 75 (0%) | 905 | 895 (-1.1%) |
| | 15% | 1 | 20 | 20 (0%) | 20 | 20 (0%) | 23 | 24 (4.35%) |
| | | 2 | 30 | 30 (0%) | 30 | 30 (0%) | 30 | 30 (0%) |
| | | 4 | 50 | 50 (0%) | 50 | 50 (0%) | 50 | 50 (0%) |

**Table 3**

| | | | Number of Readers (J) = 10 | | | | | |
|---|---|---|---|---|---|---|---|---|
| | | | Small Reader Variability | | Moderate Reader Variability | | Large Reader Variability | |
| **AUC (theta)** | **Minimum Detectable Effect Size (delta)** | **Ratio (R)** | **Reference Sample Size** | **R Output Sample Size (% Diff)** | **Reference Sample Size** | **R Output Sample Size (% Diff)** | **Reference Sample Size** | **R Output Sample Size (% Diff)** |
| **0.75** | **5%** | 1 | 116 | 116 (0%) | 201 | 200 (-0.5%) | | |
| | | 2 | 138 | 138 (0%) | 239 | 240 (0.42%) | | |
| | | 4 | 200 | 200 (0%) | 345 | 345 (0%) | | |
| | **10%** | 1 | 29 | 30 (3.45%) | 32 | 32 (0%) | 51 | 50 (-1.96%) |
| | | 2 | 35 | 36 (2.86%) | 38 | 39 (2.63%) | 60 | 60 (0%) |
| | | 4 | 50 | 50 (0%) | 55 | 55 (0%) | 87 | 90 (3.45%) |
| | **15%** | 1 | 20 | 20 (0%) | 20 | 20 (0%) | 20 | 20 (0%) |
| | | 2 | 30 | 30 (0%) | 30 | 30 (0%) | 30 | 30 (0%) |
| | | 4 | 50 | 50 (0%) | 50 | 50 (0%) | 50 | 50 (0%) |
| **0.90** | **5%** | 1 | 59 | 60 (1.69%) | 101 | 102 (0.99%) | | |
| | | 2 | 74 | 75 (1.35%) | 128 | 129 (0.78%) | | |
| | | 4 | 112 | 115 (2.68%) | 193 | 195 (1.04%) | | |
| | **10%** | 1 | 20 | 20 (0%) | 20 | 20 (0%) | 26 | 26 (0%) |
| | | 2 | 30 | 30 (0%) | 30 | 30 (0%) | 32 | 33 (3.13%) |
| | | 4 | 50 | 50 (0%) | 50 | 50 (0%) | 50 | 50 (0%) |
| | **15%** | 1 | 20 | 20 (0%) | 20 | 20 (0%) | 20 | 20 (0%) |
| | | 2 | 30 | 30 (0%) | 30 | 30 (0%) | 30 | 30 (0%) |
| | | 4 | 50 | 50 (0%) | 50 | 50 (0%) | 50 | 50 (0%) |
